# Death by scientific method: Estimated mortality associated with the failure to conduct routine prospective cumulative systematic reviews in medicine and public health

**DOI:** 10.1101/2020.10.20.20216242

**Authors:** Robert A. Hahn, Steven M. Teutsch

## Abstract

Failure to routinely assess the state of knowledge as new studies accumulate results in 1) non-use of effective interventions, 2) continued use of ineffective or harmful interventions, and 3) unnecessary research. We use a published cumulative meta-analysis of interventions to reduce the harms of acute myocardial infarctions (1966-1992), and applied population attributable risk to assess the mortality consequences of the failure to cumulatively assess the state of knowledge. Failure to use knowledge that would have been available with cumulative meta-analysis may have resulted in annual estimated mortality: 41,000 deaths from non-use of intravenous dilators, 35,000 deaths from non-use of aspirin, and 37,000 deaths annually from non-use of ß-blockers. Continued use of Class 1 anti-arrhythmic drugs, which would have been found to be harmful in 1981, resulted 39,000 deaths annually. Failure to routinely update the state knowledge can have large health consequences. The process of building knowledge and practice in medicine and public health needs fundamental revision.

## Introduction

Practitioners of medicine and public health presumably seek to act on the basis of the best available knowledge of whatever condition they are treating. They may resort to established practice or to the findings of a recent or widely cited publication.(1) Both of these approaches can be problematic. Theorists of science agree(2, 3) that the best available knowledge is ascertained by means of systematic reviews—often meta-analyses—which gather available studies of given questions (meeting specified standards), evaluate these studies (using explicit criteria), and synthesize the resulting body of evidence (with a standardized process) to determine the state of collective knowledge on the question. Systematic review can find that the evidence available is insufficient to draw a conclusion, or that available evidence of various strengths supports or negates a conclusion. The systematic review indicates the weight of available evidence to date on given quesons—a critical component in the decision of a course of action.

While use of systematic reviews for “evidence-based practice” has grown in recent decades, it is not routine, and the practice of prospective cumulative systematic reviews, i.e., adding each new study of a topic to an ongoing—thus cumulative—systematic review is rare. Nor is it clear that when such systematic evidence is available, it is readily accessible and used by practitioners.(4) In this paper we use available cumulative systematic reviews to estimate the mortality consequences of failure to routinely conduct cumulative systematic reviews and continually translate their findings into practice. Available cumulative systematic reviews examine what might have happened had ongoing systematic reviews been conducted and the findings deployed in practice.

We conclude by recommending routine prospective cumulative systematic reviews to promote those benefits. Recommendations for this practice have been made for more than 25 years,(5, 6) without substantial uptake by the research community in either medicine or public health.

## Methods

Our analysis centers on the classic study by Antman and colleagues(5) (7) which conducted retrospective cumulative meta-analyses on 15 interventions to reduce mortality among patients who had suffered acute myocardial infarctions; the study search period ran from 1966 to 1991. We assessed mortality associated with 3 interventions in the Antman study indicating benefit: intravenous vasodilators (nitroglycerin and nitroprusside) administered during hospitalization, and ß-blockers and aspirin administered both during hospitalization and for secondary prevention after hospital discharge. We also assessed the consequences of one intervention shown by cumulative meta-analysis to be harmful for routine use in the treatment of acute myocardial infarctions—Class 1 anti-arrhythmic drugs; because this harm was unrecognized, the harmful practice continued.(5)

Antman’s study provides three critical pieces of information:

1. the year in which the ongoing, cumulative meta analysis first indicated benefit (or harm) at a chosen level of statistical significance
2. the effect size determined—commonly an odds ratio indicating the proportion of mortality reduced by the intervention compared with placebo or no intervention, and
3. the year in which the intervention became routine in practice (or was eliminated in practice because it is harmful or ineffective).

Routine practice was assessed by examination of samples of published review papers and texts on treatment for myocardial infarction in each year over recent decades. Review papers and textbook statements were classified as: a) recommending routine use, b) recommending use in specific circumstances, c) recommending rare use or nonuse, d) experimental, or e) use not mentioned.

We used information about mortality from acute myocardial infarction before, during, and after hospitalization to estimate mortality associated with failure to use information on benefits (or harms) of interventions that were not used (or were used) because cumulative systematic reviews had not been conducted, so that the requisite knowledge was not available. Deaths from myocardial infarction occurring prior to hospitalization were excluded from the analysis because they would not have been subject to the interventions under study. Deaths that occurred after hospital admission and after hospital discharge were included because they might have been addressed by the interventions for either acute or longer-term treatment. While rates and numbers of deaths associated with myocardial infarction have changed over the study period, we use the number of deaths at the approximate midpoint of the study period, i.e., 1980 to estimate attributable mortality.

We use the epidemiological method of population attributable risk (PAR) to estimate the number of deaths that might have been averted had the intervention discovered retrospectively by cumu^-^ lative meta analysis been known.

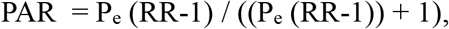

where P_e_ is the prevalence of nonuse of the practice we are assessing, and RR is the relative risk of the outcome, i.e., death in this case, associated with nonuse of the practice, compared with use, i.e., the number of deaths that might have been averted with use of the preventive measure in question. We assess the consequences of 100% prevalence of nonuse of the preventive measure. Thus, P_e_ drops from the equation, and the equation becomes simply PAR = (RR-1) / RR. In sensitivity analyses, we assess the benefits of partial adoption, i.e., P_e_ <100%, or changing other parameters. The RR is the inverse of the odds ratio associated with the benefit of each intervention explored and reported in Lau.(7)

## Results and Discussion

### Mortality associated with myocardial infarction (MI)

We used an estimate of the number of MI patients in the population with an acute myocardial infarction (ICD code 410) in 1980 from the CDC’s National Hospital Discharge Survey (https://www.cdc.gov/nchs/nhds/index.htm). We use fatality rates for first and subsequent MIs from a synthesis of estimates of pre-hospital, in-hospital, and post-hospital mortality rates associated with an acute MI (Table 1)(8) to reconstruct the number of first and subsequent MIs in-1980, the number of in-hospital deaths, and the number of deaths within 3 years of hospital discharge from hospital survivors.

**Table 1.** Estimating MI mortality rates Pre-, In-, and Post-Hospitalization (Law 2002)

| **First or subsequent MI** | Pre hospital fatality | In-Hospital case fatality | Fatality 1st year Post-Hospital | Fatality after first year |
| --- | --- | --- | --- | --- |
| **First** | 0.23 | 0.13 | 0.1 | 0.05 |
| **Subsequent** | 0.33 | 0.2 | 0.2 | 0.1 |

We assume that the reporting of ICD 410 is for first MI, and we reconstruct the number of subsequent incident MIs using proportions from other sources.(https://www.cdc.gov/heartdisease/facts.htm) In a sensitivity analysis, we assume ICD 410 reporting to include both first and subsequent MIs. Approximately 225,000 MI patients died before reaching the hospital, i.e., 49% of the deaths assessed in this analysis (Table 2). There were an estimated 95,000 in-hospital deaths (21%) and 137,000 (30%) after-hospital deaths in 1980 (Table 2).

**Table 2.** Estimating In- and Post-Hospital MI Total Deaths in 1980

| Year | Estimated MIs in US | pre hospital death (% incidence) | pre hospital death | Hospital admission | Hospital case fatality | In-Hospital deaths | MI patients discharged alive | Mortality/yr 1st year | Mortality/after first year | Deaths post discharge —3 years | Total deaths acute MI. In and post hospital 3 yrs |
| --- | --- | --- | --- | --- | --- | --- | --- | --- | --- | --- | --- |
| **First acute MI, 1980** | 559,740 | 0.23 | 128,740 | 431,000 | 0.13 | 56,030 | 374,970 | 0.10 | 0.05 | 74,994 | 131,024 |
| **Subsequent acute MI, 1980** | 290,222 | 0.33 | 95,773 | 194,449 | 0.2 | 38,890 | 155,559 | 0.2 | 0.1 | 62,224 | 101,113 |
| **Total acute MI, 1980** | 849,962 |  | 224,514 | 625,449 |  | 94,920 | 530,529 |  |  | 137,218 | 232,137 |

### Estimating mortality attributable to nonuse of interventions which were not known, but could have been known

In the hospital setting, the annual number of deaths associated with the non-use of interventions that would have been available had cumulative meta-analyses been conducted range from 12,000 for the non-use of intravenous or oral ß-blockers to 41,000 for the non-use of intravenous vasodilators (Table 3).

**Table 3.** Increased deaths/year associated with non-use of selected interventions to reduce mortality associated with myocardial infarction.

| Intervention | yr meta anal finding | yr routine use | gap years | final OR | RR death with intervention nonuse | PAR % | Increased deaths/yr with intervention nonuse. In Hospital Deaths |
| --- | --- | --- | --- | --- | --- | --- | --- |
| **A. MORTALITY IN ACUTE MI IN HOSPITAL** |  |  |  |  |  | deaths associated with MI, in hospital | 94,920 |
| intravenous vasodilators (nitroglycerin and nitroprusside) | 1981 (<0.05) | 1990 | 9 | 0.57 (0.41 - 0.79) | 1.75 | 0.43 | 40,680 |
| aspirin | 1988 (<0.00001) | 1990 | 2 | 0.77 (0.70 - 0.84) | 1.3 | 0.23 | 21,905 |
| intravenous or oral β-blockers | 1986 (<0.05) | 1991 | 5 | 0.88 (0.80 - 0.98) | 1.14 | 0.12 | 11,657 |
| **B. MORTALITY FOLLOWING ACUTE MI (SECONDARY PREVENTION) AFTER HOSPITAL DISCHARGE** |  |  |  |  |  |  | Increased deaths/yr with intervention nonuse. After Hospital Deaths—3 yrs |
|  |  |  |  |  |  | After hospital deaths | 137,218 |
| antiplatelet drugs/ASPIRIN | 1981 (<0.05) |  |  | 0.90 (0.82 - 1.00) | 1.11 | 0.10 | 13,598 |
| oral β-blockers, | 1977 (<0.05) | 1983 | 6 | 0.81 (0.73 - 0.89) | 1.23 | 0.19 | 25,659 |
| type I antiarrhythmic drugs | 1981 (<0.05) |  |  | 1.28 (1.02 - 1.61) | 0.78 | -0.28 | -38,703 |

Table 3. Annual deaths attributable to non use of interventions for intervention for acute MI in A. Acute treatment and B. 3 years post-discharge.

In the post-hospital setting, failure to use secondary preventive measures that would have been available had cumulative meta-analyses been conducted are 14,000 for the non-use of antiplatelet drugs (predominantly aspirin) and 26,000 for the non-use of oral ß-blockers (Table 3). The use of type I antiarrhythmic drugs in secondary prevention was found to be harmful, with a summary odds ratio of 1.28 (Table 3). Their routine use can be estimated to be associated with 39,000 deaths per year (Table 3).

### Sensitivity analyses

It is likely that at least some of the parameters used in the analysis are imprecise. We modify several of the parameters in our analysis to determine effects on PAR and estimated deaths We use the example of aspirin use. First we ask how many deaths would occur if use were 50% instead of 0%; the number of deaths in-and after-hospitalization are kept constant(Table 4). Annual in-hospital deaths would be approximately 12,000, and after-hospital deaths would be approximately 7,000 (Table 4). Then we ask what happens if all MIs are reported in ICD code 410, including both first and subsequent MIs; in this case, the PAR remains constant, but the number of deaths changes (Table 4). In this case, annual in-hospital deaths would be approximately 13,000, and after-hospital deaths would be approximately 1,000 (Table 4). Finally, we ask what would happen were the effect sizes reduced by 50%, i.e., to 1.15 for acute treatment and 1.055 for post-discharge treatment. The result would be reductions in mortality equivalent to increasing the use of the intervention to 50% (Table 4).

The mortality costs of the failure to routinely conduct prospective cumulative systematic reviews at the conclusion of each new study are very high. Even when underlying parameters are re^-^ duced in sensitivity analyses, the number of annual deaths remains unacceptably high. There are other costs as well—the financial, opportunity, and human costs of continued randomized trials when the basic question has already been answered, and the moral cost of not deploying the best possible treatment for given conditions.

**Table 4.** Sensitivity Analyses for Aspirin Use In- and After-Hospitalization

| Modification | In-hospital attributable deaths |  | After-hospital attributable deaths |  |
| --- | --- | --- | --- | --- |
| Intervention used for 50% of cases, i.e., $P_e = 0.5$ | 0.13 | 12,340 | 0.05 | 7,153 |
| All MIs reported in first MI category, i.e., no “subsequent MI” | 0.23 | 12,887 | 0.10 | 1,360 |
| Effect size 50% of that reported, i.e., 1.15 for acute treatment, 1.055 for post-discharge treatment | 0.13 | 12,340 | 0.05 | 7,153 |

The data sources for the present analysis are less than optimal. As Feinlieb, then Director of the National Center for Health Statistics, noted in 1984, “Unfortunately, this country has no method or system for standardized complete reporting of new myocardial infarctions and no incidence data representative of the national population”(9)—a situation that has not changed. Estimates of population incidence of MIs (i.e., ICD 410) range between 450,000 and 600,000, indicating that the estimate we use as the basis for our PAR findings is consistent with other reports.(9, 10) when the basic question has already been answered, and the moral cost of not deploying the bestpossible treatment for given conditions.

The data sources for the present analysis are less than optimal. As Feinlieb, then Director of theNational Center for Health Statistics, noted in 1984, “Unfortunately, this country hasno method or systemfor standardized complete reporting of newmyocardial infarctions and no incidencedatarepresentative of thenational population"(9)—a situation that has not changed. Estimatesof population incidence of MIs (i.e., ICD 410) range between 450,000 and 600,000, indicating that the estimate we use as the basis for our PAR findings is consistent with other reports.(9, 10) (https://www.cdc.gov/heartdisease/facts.htm). Rates of intervention use are not known, and es-timates of rates of MI fatality before, during, and after hospitalization may also be imprecise. Sensitivity analyses with increased intervention use and reduced estimates of MI incidence stillyield substantial harm from failure to conduct prospective cumulative systematic reviews.

We are aware of two cumulative analyses of public health interventions—one on infant sleep po-sitions to avoid Sudden Infant Death Syndrome (SIDS)(11) and one on passive smoking.(12) Gilbert and colleagues report that prone sleeping for infants was recommended in books between 1943 and 1988. Gilbert and colleagues also find that by 1970, cumulative meta-analyses would have indicated increased SIDS mortality associated with prone sleeping, confirmed by later data. They estimate that approximately 60,000 infant deaths would have been averted had this knowl-edge been applied.(11) Rates of intervention use are not known, and estimates of rates of MI fatality before, during, and after hospitalization may also be imprecise. Sensitivity analyses with increased intervention use and reduced estimates of MI incidence still yield substantial harm from failure to conduct prospective cumulative systematic reviews.

We are aware of two cumulative analyses of public health interventions—one on infant sleep positions to avoid Sudden Infant Death Syndrome (SIDS)(11) and one on passive smoking.(12) Gilbert and colleagues report that prone sleeping for infants was recommended in books between 1943 and 1988. Gilbert and colleagues also find that by 1970, cumulative meta-analyses would have indicated increased SIDS mortality associated with prone sleeping, confirmed by later data. They estimate that approximately 60,000 infant deaths would have been averted had this knowledge been applied.(11)

In another domain, Taylor and colleagues(12) find that meta-analytic evidence for the harms of secondhand smoke from cigarette smokers could have been determined in 1972. However, a causal link was only recognized in the Surgeon General’s Report on Smoking in 1986.(13) It is estimated that secondhand smoke from cigarette smokers kills 41,000 adults per year in the U.S. Indoor air smoking policies began to expand across the nation in about 1984.(14) Levy and colleagues(1) (2018) estimate that an average of 773,000 deaths per year were averted globally between 2007 and 2014 by smoke free environmental policies.

The institutionalization of prospective cumulative systematic reviews is not a simple matter and is likely to require institutional change and global coordination. Elements of the process would require a registry of ongoing research on different topics and a mechanism/process for updating the current cumulative meta-analysis with new findings. A structure would have to be stood up to manage and coordinate such efforts. Details would have to be established, e.g., determining the level of finding certainty at which to decide that evidence was sufficient.(15) Such registries could also serve as foundations for continuing research in that they indicate issues meriting exploration.

## Conclusions

Ongoing cumulative meta-analysis should be routine in public health. The process of building knowledge and practice in medicine and public health needs fundamental revision. The benefits of such a process could be the prevention of hundreds of thousands of deaths each year.

## Data Availability

available from author

